# Attributable mortality of vancomycin resistance in ampicillin-resistant *Enterococcus faecium* bacteremia in Denmark and the Netherlands: a matched cohort study

**DOI:** 10.1101/2020.05.10.20049783

**Authors:** Wouter C. Rottier, Mette Pinholt, Akke K. van der Bij, Magnus Arpi, Sybrandus N. Blank, Marrigje H. Nabuurs-Franssen, Gijs J.H.M. Ruijs, Matthijs Tersmette, Jacobus M. Ossewaarde, Rolf H. Groenwold, Henrik Westh, Marc J.M. Bonten, on behalf of the Dutch VRE Bacteremia Investigators and the Danish Collaborative Bacteraemia Network (DACOBAN)

**Author notes:** Department of Medical Microbiology and Immunology, Diakonessenhuis, Utrecht, the Netherlands. Authors contributed equally. **Corresponding author:** Wouter C. Rottier, Julius Center for Health Sciences and Primary Care, University Medical Center Utrecht, Huispostnummer STR 6.131, Postbus 85500, 3508 GA Utrecht, the Netherlands.

## Abstract

**Background:** In many European hospitals, ampicillin-resistant *Enterococcus faecium* (ARE) is endemic, while outbreaks of vancomycin-resistant *E. faecium* (VRE), belonging to the same genetic lineage, are increasingly reported. We studied the attributable mortality due to vancomycin resistance in patients with *E. faecium* bacteremia and evaluated whether this is mediated by a delay in appropriate antibiotic therapy.

**Methods:** In a retrospective matched cohort study, patients with VRE bacteremia occurring between 2009 and 2014 in 20 Dutch and Danish hospitals were matched to patients with ARE bacteremia, on hospital, ward, length of hospital stay prior to bacteremia, and age. The risk ratio (RR) for 30-day mortality contrasting VRE with ARE was estimated with further analytic control for confounding factors.

**Results:** In all, 63 VRE and 234 ARE episodes were matched (36 and 130 for the Netherlands and 27 and 104 for Denmark). Crude 30-day mortality was 27% and 38% for ARE in the Netherlands and Denmark, respectively, and 33% and 48% for VRE in the respective countries. The adjusted RR for 30-day mortality for VRE was 1.54 (95% confidence interval (CI) 1.06-2.25). Although appropriate therapy was initiated later for VRE than for ARE bacteremia, this did not appear to mediate the increased mortality risk.

**Conclusions:** Compared to ARE bacteremia, VRE bacteremia was associated with higher 30-day mortality. One explanation for this association would be increased virulence of VRE, although both phenotypes belong to the same well-characterized core genomic lineage. Alternatively, it may be the result of unmeasured confounding.

## Introduction

As many other countries, the Netherlands and Denmark have faced increasingly frequent polyclonal hospital outbreaks of *Enterococcus faecium* with combined resistance to ampicillin and vancomycin (VRE) during the past years [1,2]. In these countries, ampicillin-resistant, vancomycin-susceptible *E. faecium* (ARE) has become the dominant hospital phenotype of *E. faecium* in recent decades [2,3], Since hospital-acquired VRE and ARE are genetically indistinguishable at the core genome level, VRE is assumed to have originated from the omnipresent ARE through acquisition of *vanA* or *vanB* genes [4–7], In both countries, infection control policies have been implemented to prevent nosocomial transmission of VRE (contact precautions for VRE carriers, supplemented by contact tracing and augmented general hygiene measures in case of outbreaks [8,9]), but not for ARE. Failure to control VRE transmission will most likely result in VRE endemicity, because the nosocomial ARE populations will in part be supplanted by VRE [3,10],

Controlling VRE outbreaks imposes a great burden on finances and hospital personnel [11]. To make an appropriate cost-benefit analysis of containing VRE spread in a healthcare system, it is essential to quantify the benefits of such a strategy. The most important threat for individual patients is the adversity patients will experience due to VRE infection as compared to ARE infection. A meta-analysis reported increased mortality after VRE bacteremia compared to ARE bacteremia [12], but most studies included had been performed before effective antibiotics for VRE were available. Since then, few have attempted to quantify the effects of VRE infection compared to ARE infection, and those available often suffered from methodological drawbacks, such as combining *E. faecium* and *Enterococcus faecalis* infection and incomplete control for confounding [13].

We, therefore, sought to investigate the fraction of mortality in VRE bacteremia superimposed by vancomycin resistance, in both the Netherlands and Denmark. We also analyzed whether any such increase is the result of a delay in appropriate antibiotic therapy, as this is *a priori* the most likely mediating mechanism.

## Methods

### Study design, setting and participants

We addressed a causal research question with an observational study in which confounding bias was dealt with in a two-stepped approach. First, by means of matching, patients with ARE bacteremia and with underlying disease severity similar to the VRE bacteremia patients were chosen as comparison group. Second, we controlled for remaining imbalances in confounding factors after matching by means of adjustment in multivariable models.

This resulted in a retrospective matched cohort study in which episodes of bacteremia caused by *E. faecium* with co-resistance to ampicillin and vancomycin (designated as VRE) were compared to control episodes of bacteremia caused by *E. faecium* with resistance to ampicillin and susceptibility to vancomycin (designated as ARE). Episodes with ampicillin-susceptible VRE bacteremias were excluded. Depending on the availability, a maximum of 4 ARE bacteremias were matched to each VRE bacteremia, using the variables hospital, hospital ward at bacteremia onset, age and length of stay prior to bacteremia (see Supplementary Material for a complete description).

No formal sample size calculation was performed, as in both involved countries, VRE bacteremias are rare occurrences, and we had to rely on the willingness of hospitals country-wide to participate. In the Netherlands, VRE bacteremia episodes were identified in 13 hospitals through the national surveillance system ISIS-AR [14], and eleven participated in this study, as did five hospitals not linked to ISIS-AR (see Supplementary Table 1 for details of participating hospitals). In Denmark, the DACOBAN database was used to identify patients with VRE bacteremia. DACOBAN is a registry of all positive blood cultures from 10 of 11 hospitals in the Capital Region of Denmark (the exception being the tertiary referral center Rigshospitalet) [15]. Patients with VRE bacteremia were identified in five hospitals of which four participated in this study.

In the Netherlands, we included patients with VRE bacteremia that occurred between 1 January 2009 and 1 January 2013, with deviations in some hospitals (Supplementary Table 1). In Denmark, we included patients with VRE bacteremia that occurred between 1 January 2012 and 1 January 2015.

Minimum inhibitory concentrations (MICs) for ampicillin and vancomycin were used as reported by local laboratories. All Danish laboratories interpreted antimicrobial susceptibility according to EUCAST standards, but most Dutch laboratories switched from CLSI to EUCAST standards in recent years [14]. Vancomycin resistance had to be confirmed by E-test or demonstration of the presence of *vanA* or *vanB*. The specific VRE genotype was based on PCR-testing or on teicoplanin susceptibility (resistant categorized as *vanA*, susceptible as *vanB*) if PCR testing had not been performed.

The Institutional Review Board of the coordinating center judged the study to be exempt from the Dutch Medical Research Involving Human Subjects Law due to its retrospective nature. Informed consent was not necessary, as data were provided anonymized by treating physicians. In all participating study sites, local regulations for such studies were followed. In Denmark, the study was approved by the Danish Data Protection Agency (registered under 2012-58-0004) and the Danish Health and Medicines Authority (registered under 3-3013-1118/1).

### Data collection

After selection of cases and controls, charts were manually reviewed with the date of the index blood culture (bacteremia onset) as reference date. A description of the potential confounding variables and infection-related variables for which data were collected is provided in the Supplementary Material.

Additionally, antibiotic use was registered from 30 days prior to bacteremia onset until 14 days after onset, including type of antibiotic, route of administration, and starting and stopping dates. Antibiotic use prior to bacteremia was considered a potential confounder, whereas treatment provided for the *E. faecium* bacteremia episode was considered the main intermediate variable on the causal pathway leading from vancomycin resistance to increased mortality. To analyze this variable, on each calendar day from bacteremia onset onwards (considered day 0), antibiotic treatment was categorized as (a) either *E. faecium*-covering (i.e. including vancomycin, linezolid, daptomycin, teicoplanin, quinupristin/dalfopristin and/or tigecyclin, regardless of vancomycin resistance phenotype) or not, and (b) appropriate (i.e. all of the aforementioned antibiotics for ARE infection, all except vancomycin for *vanB* VRE infection, and all except vancomycin and teicoplanin for *vanA* VRE infection) or inappropriate.

The primary outcome of the study was mortality within 30 days of bacteremia onset, and secondary outcomes were mortality within 1 year, in-hospital mortality, length of hospital stay after bacteremia onset, and intensive care unit admission within 7 days of bacteremia onset. For all bacteremia cases, follow-up data (censoring date or date of death) for at least 30 days after bacteremia onset, but preferably up to 1 year after bacteremia onset were collected.

### Statistical analysis

The relation between ARE/VRE and 30-day mortality was estimated using Cox regression models, unadjusted as well as adjusted for potential confounding variables. All models were Cox proportional hazards models, with stratification on matched sets, robust standard errors, and correlation between individuals that were included multiple times. For models without censoring, all episodes were given the same arbitrary follow-up time and the Efron approximation for tied survival times was used, so that hazard ratios (HR) could be interpreted as risk ratios (RR) [16]. The standard adjusted models involved inclusion of all potential confounders a priori deemed relevant by us to achieve optimal correction, followed by removal of redundant variables to increase precision [17]. As a sensitivity analysis, stepwise addition and removal of potential confounders was performed, starting from a model including only the exposure of interest. In the Supplementary Material, exact procedures are described. In case of missing values for the variables included in a model, individuals were excluded from the analysis. Several additional models were created to evaluate mediation of the effect of VRE on mortality through appropriateness of therapy. For this, an interaction between vancomycin resistance and appropriateness of therapy was included. As appropriateness of therapy is a time-varying variable, three models were created in which the baseline was moved to the end of day 0 (day of the index blood culture), +1, and +2, respectively. Patients having died or censored before or on the day of the baseline were removed from the analysis. Appropriateness of therapy in each model reflected the state at baseline.

All statistical analyses were performed in R (version 3.6.1) [18], with the use of packages *survival* [19], *cmprsk* [20], *rms* [21], *mice* [22] and *xtable* [23],

## Results

### Patient characteristics

In all, 63 VRE episodes were matched to 234 ARE episodes (36 and 130 for the Netherlands, and 27 and 104 for Denmark). VRE and matched ARE bacteremia episodes had largely similar characteristics (Table 1 and Supplementary Table 2). Differences between both countries were also present, most prominently involving treatment restriction prior to bacteremia. The latter variable is generally registered on a dedicated location in Dutch health records, but had to be abstracted from written notes in Denmark. Also, comorbidities were retrieved from the DACOBAN registry in Denmark, whereas they were abstracted from medical notes in the Netherlands.

**Table 1.** Characteristics and outcomes of VRE and matched ARE bacteremias

|  | Netherlands |  | Denmark |  |
| --- | --- | --- | --- | --- |
|  | ARE bacteremia,<br>n/N with data<br>(%) | VRE bacteremia,<br>n/N with data<br>(%) | ARE bacteremia,<br>n/N with data<br>(%) | VRE bacteremia,<br>n/N with data<br>(%) |
| **Potential confounding variables** |  |  |  |  |
| Female | 47/130 (36) | 19/36 (53) | 51/104 (49) | 10/27 (37) |
| Age, median (IQR) | 70 (62-76) | 69 (62-76) | 69 (63-77) | 71 (58-76) |
| Hospital ward at bacteremia onset |  |  |  |  |
| - Internal medicine | 47/130 (36) | 15/36 (42) | 31/104 (30) | 8/27 (30) |
| - ICU | 48/130 (37) | 11/36 (31) | 25/104 (24) | 6/27 (22) |
| - Gastro-enterology/surgery | 34/130 (26) | 9/36 (25) | 31/104 (30) | 8/27 (30) |
| - Other | 1/130 (1) | 1/36 (3) | 17/104 (16) | 5/27 (19) |
| Bacteremia origin |  |  |  |  |
| - Hospital-onset | 113/130 (87) | 29/36 (81) | 93/104 (89) | 24/27 (89) |
| - Healthcare-associated | 15/130 (12) | 4/36 (11) | 10/104 (10) | 1/27 (4) |
| - Community-onset | 2/130 (2) | 3/36 (8) | 1/104 (1) | 1/27 (4) |
| Length of hospital stay prior to bacteremia, median (IQR) | 17 (11-24) | 20 (14-36) | 18 (6-24) | 21 (10-29) |
| Preceding hospital admission within 3 months prior to bacteremia | 58/130 (45) | 15/36 (42) | 47/103 (46) | 13/26 (50) |
| Charlson index, median (IQR) | 2 (2-3) | 2 (1-4) | 3 (1-4) | 3 (2-6) |
| Hematological malignancy - under treatment | 31/130 (24) | 10/36 (28) | 9/104 (9) | 6/27 (22) |
| Metastasized solid malignancy | 13/130 (10) | 1/36 (3) | 13/104 (12) | 6/27 (22) |
| Neutropenia at bacteremia onset | 32/130 (25) | 11/36 (31) | 8/104 (8) | 6/27 (22) |
| Treatment restriction in place at bacteremia onset | 26/130 (20) | 10/36 (28) | 5/102 (5) | 2/27 (7) |
| Surgical procedure within 30 days prior to bacteremia | 46/130 (35) | 9/36 (25) | 33/103 (32) | 12/27 (44) |
| Known colonization with *E. faecium* |  |  |  |  |
| - No | 90/130 (69) | 18/36 (50) | 88/104 (85) | 16/27 (59) |
| - Yes: ARE | 38/130 (29) | 7/36 (19) | 16/104 (15) | 2/27 (7) |
| - Yes: VRE | 2/130 (2) | 11/36 (31) | 0/104 (0) | 9/27 (33) |
| Antibiotic use within 30 days prior to bacteremia | 122/130 (94) | 33/36 (92) | 95/104 (91) | 25/27 (93) |
| Vancomycin use within 30 days prior to bacteremia | 13/130 (10) | 15/36 (42) | 9/104 (9) | 6/27 (22) |
| **Infection-related variables** |  |  |  |  |
| Polymicrobial bacteremia | 35/130 (27) | 8/36 (22) | 31/103 (30) | 11/27 (41) |
| Severe sepsis at bacteremia onset | 30/129 (23) | 10/36 (28) | 20/104 (19) | 4/27 (15) |
| Bacteremia source |  |  |  |  |
| - Primary bacteremia/central line infection/not identifiable from medical file | 60/130 (46) | 19/36 (53) | 59/104 (57) | 11/27 (41) |
| - Biliary tract infection | 15/130 (12) | 4/36 (11) | 10/104 (10) | 2/27 (7) |
| - Other intra-abdominal infection | 35/130 (27) | 9/36 (25) | 12/104 (12) | 7/27 (26) |
| - Other | 20/130 (15) | 4/36 (11) | 23/104 (22) | 7/27 (26) |
| Source control performed before day +7<sup>a</sup> (if applicable to source) | 38/79 (48) | 8/19 (42) | 25/47 (53) | 4/13 (31) |
| **Outcome variables** |  |  |  |  |
| ICU admission before day +7<sup>a</sup> (if not yet in ICU) | 4/82 (5) | 4/25 (16) | 9/79 (11) | 1/21 (5) |
| Length of hospital stay after bacteremia onset (median, IQR) | 22 (10-38) | 13 (8-24) | 16 (8-36) | 14 (6-30) |
| In-hospital mortality | 34/130 (26) | 10/36 (28) | 38/104 (37) | 16/27 (59) |
| Mortality before day +30<sup>a</sup> | 35/130 (27) | 12/36 (33) | 40/104 (38) | 13/27 (48) |

This table presents a selection of recorded variables. A full overview is available in Supplementary Table 2.
Abbreviations: ICU, intensive care unit; IQR, interquartile range.
<sup>a</sup> Day 0 is the day of the index blood culture of the ARE/VRE bacteremia episode.

Most VRE were *vanA* (n = 41, 65%), 19 were *vanB* (30%), one isolate carried both *vanA* and *vanB* and two isolates could not be categorized. All VRE isolates from Denmark (n = 27) were *vanA*. Seventeen isolates were categorized based on teicoplanin susceptibility (3 *vanA* and 14 *vanB*, all from the Netherlands).

### Mortality

All patients could be assessed for 30-day mortality, and 76% of censored patients had a follow-up time of at least one year. Crude 30-day mortality was 40% for VRE and 32% for ARE: 33% and 27% for VRE and ARE, respectively, in the Netherlands and 48% and 38% in Denmark. In the Netherlands, 30-day mortality per VRE phenotype was 29% for *vanA* and 37% for *vanB*. The unadjusted RR for 30-day mortality of VRE (compared to ARE) was 1.27 (95% confidence interval (CI) 0.87-1.84; 1.16 (95% CI 0.65-2.06) for the Netherlands, 1.37 (95% CI 0.84-2.25) for Denmark; Table 2). Adjustment for confounding increased the RR to 1.54 (95% CI 1.06-2.25). Within the Dutch subgroup, addition of the confounder *Acute Physiology Score before bacteremia onset* to an otherwise optimally adjusted model reduced the RR of vancomycin resistance from 1.62 to 1.17 (see Supplementary Material).

**Table 2.** Regression models for 30-day mortality

|  | Unadjusted models |  |  | Combined adjusted models |  |  |
| --- | --- | --- | --- | --- | --- | --- |
|  | Combined | Netherlands | Denmark | Main analysis | Sensitivity analysis | Therapy added |
| *No of observations* | 297 | 166 | 131 | 297 | 295 | 297 |
| *No of events* | 100 | 47 | 53 | 100 | 99 | 100 |
| *No of variables* | 1 | 1 | 1 | 5 | 8 | 6 |
| *Events to variables* | 100 | 47 | 53 | 20 | 12.4 | 16.7 |
| Vancomycin resistance | 1.27 (0.87-1.84) | 1.16 (0.65-2.06) | 1.37 (0.84-2.25) | 1.54 (1.06-2.25) | 1.49 (0.99-2.22) | 1.55 (1.07-2.26) |
| Inappropriate therapy on day of index blood culture |  |  |  |  |  | 2.79 (0.99-7.86) |

This table presents risk ratios (RR, with 95% CI) for 30-day mortality for the exposure of interest (*vancomycin resistance*), with and without adjustment for confounding variables, and with and without the intermediate variable *inappropriate therapy on day of index blood culture*. In the Supplementary Material, the adjustment procedure is described. Combined models include data from both the Netherlands and Denmark.

One year mortality was larger among Danish patients with VRE compared to patients with ARE bacteremia or Dutch patients with VRE or ARE bacteremia (Figure 1). In the multivariable Cox model for mortality up to one year, the HR for VRE amounted to 1.25 (95% CI 0.80-1.98; Table 3).

**Figure 1.**
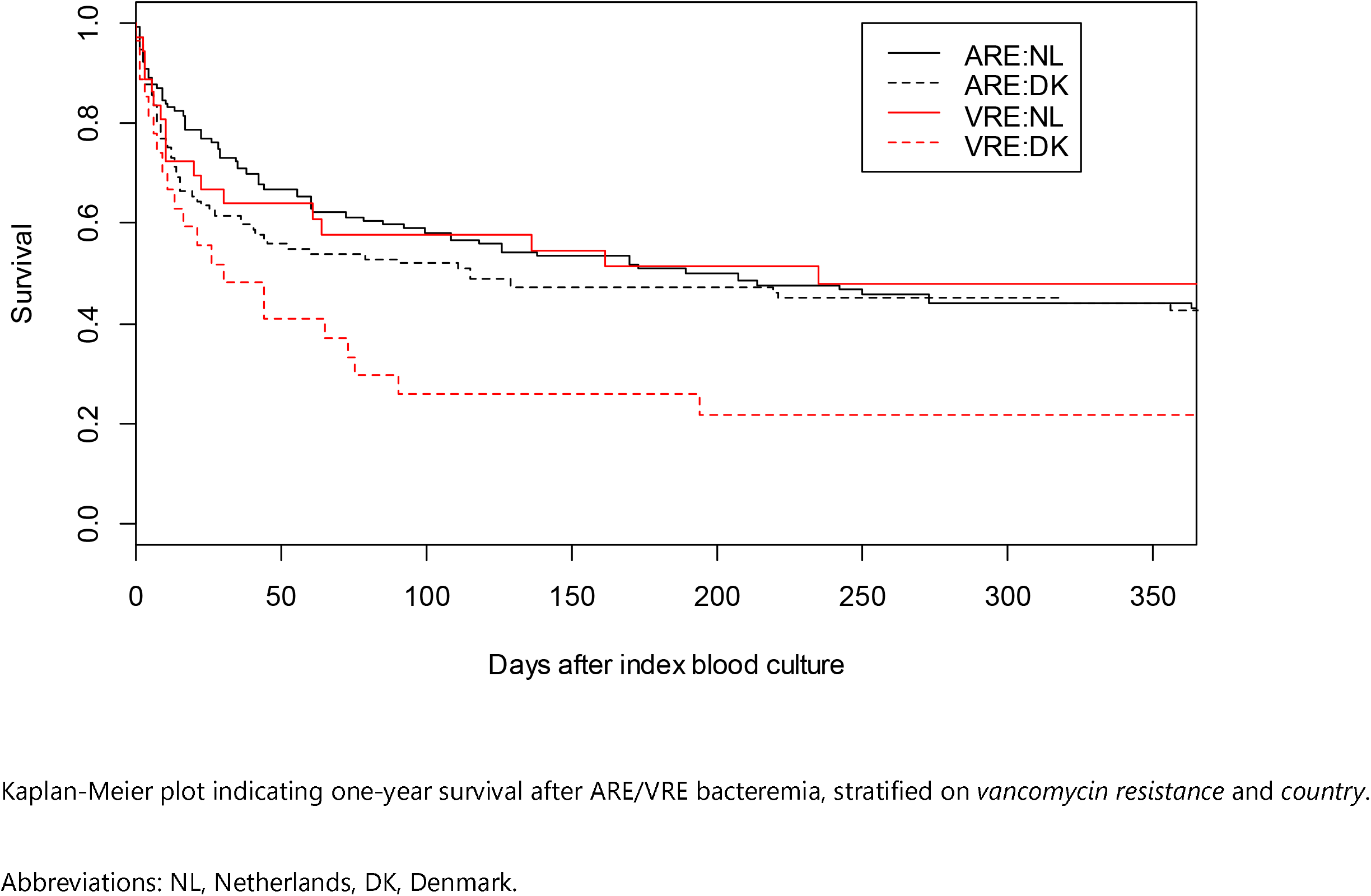
One-year survival curve for VRE and matched ARE bacteremias

**Table 3.** Regression models for one-year follow-up for mortality

|  | Unadjusted models |  |  | Combined adjusted models |  |
| --- | --- | --- | --- | --- | --- |
|  | Combined | Netherlands | Denmark | Main analysis | Sensitivity analysis |
| *No of observations* | 297 | 166 | 131 | 294 | 297 |
| *No of events* | 170 | 90 | 80 | 170 | 170 |
| *No of variables* | 1 | 1 | 1 | 12 | 7 |
| *Events to variables* | 170 | 90 | 80 | 14.2 | 24.3 |
| Vancomycin resistance | 1.18 (0.84-1.65) | 0.91 (0.53-1.55) | 1.51 (0.97-2.36) | 1.25 (0.80-1.98) | 1.46 (0.95-2.25) |

This table presents hazard ratios (HR, with 95% CI) for mortality (follow-up of 1 year with censoring) for the exposure of interest (*vancomycin resistance*), with and without adjustment for confounding variables. In the Supplementary Material, the adjustment procedure is described. Combined models include data from both the Netherlands and Denmark.

### Antibiotic therapy

Visual inspection of cumulative incidence plots revealed that initiation of *E. faecium*-covering antibiotic therapy occurred faster in VRE than in ARE episodes (Figure 2), but that initiation of appropriate antibiotic therapy occurred faster for ARE compared to VRE bacteremia (Figure 3). In Denmark, appropriate antibiotic therapy for both ARE and VRE bacteremia was started earlier than in the Netherlands, and often consisted of linezolid daptomycin combination treatment (Table 4).

**Figure 2.**
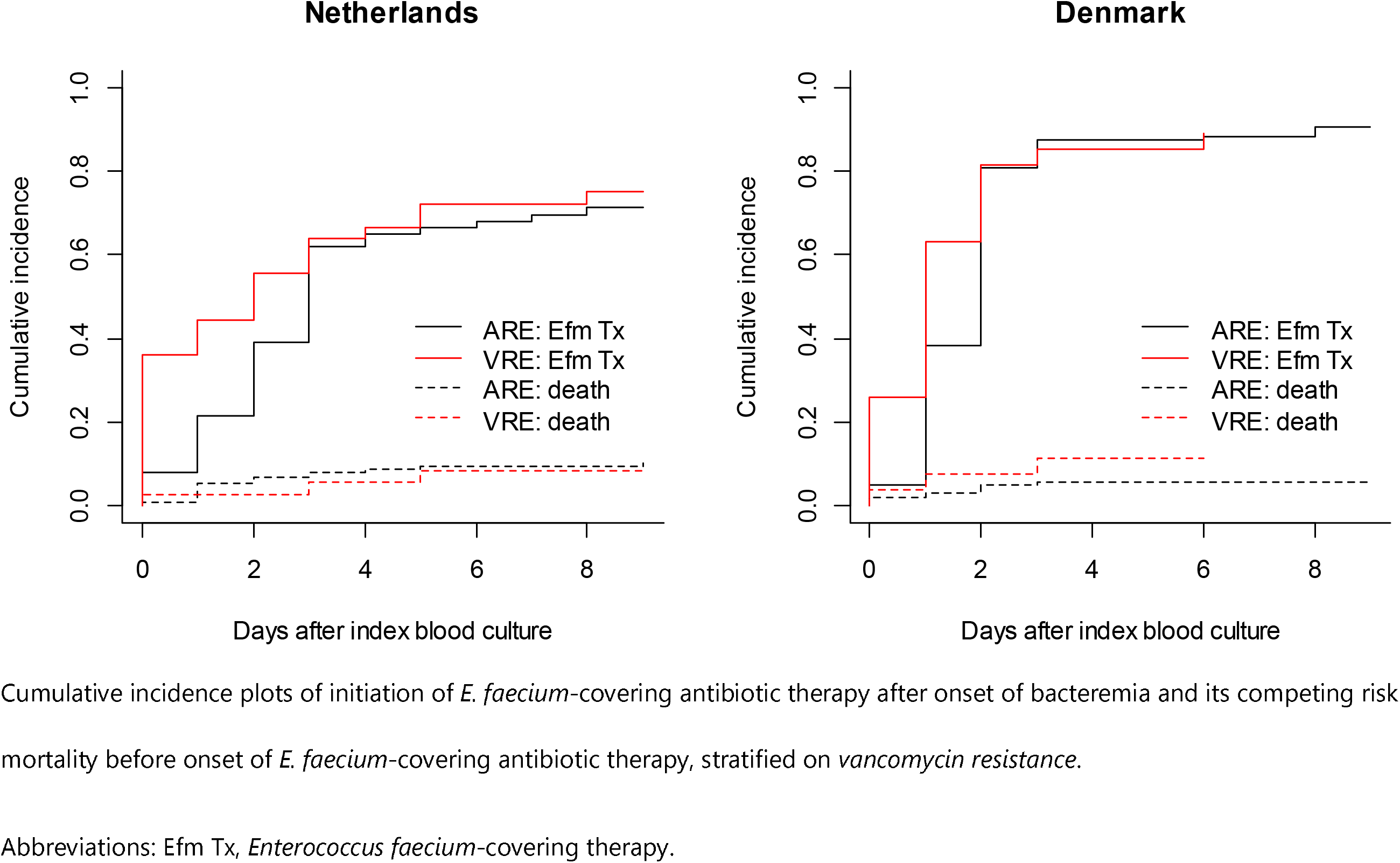
Time to *Enterococcus faecium*-covering antibiotic therapy for VRE and matched ARE bacteremias

**Figure 3.**
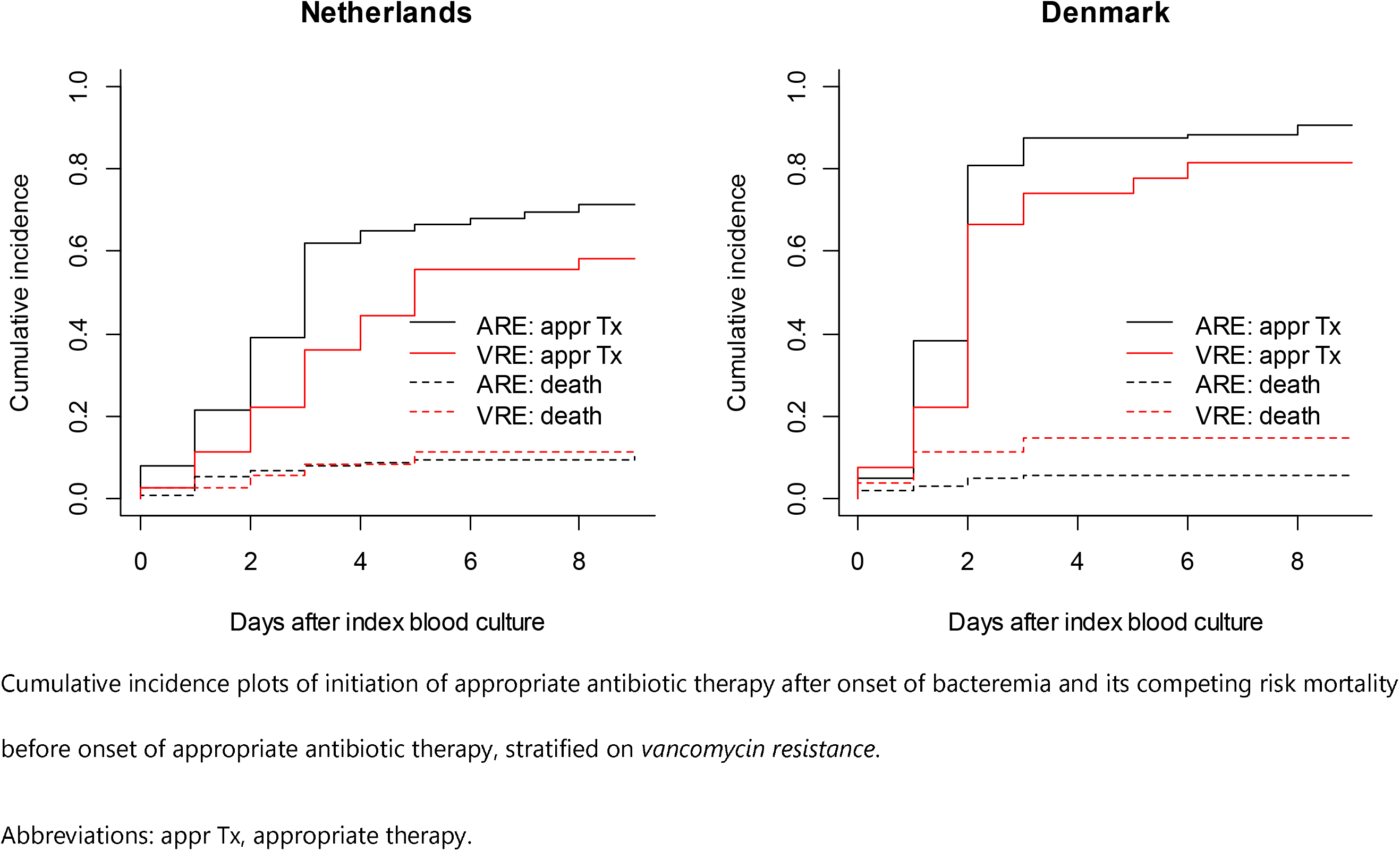
Time to appropriate antibiotic therapy for VRE and matched ARE bacteremias

**Table 4.** Overview of antibiotic therapy for VRE and matched ARE bacteremias

|  | End of day 0<sup>a</sup> |  |  |  | End of day +3<sup>a</sup> |  |  |  |
| --- | --- | --- | --- | --- | --- | --- | --- | --- |
|  | ARE |  | VRE |  | ARE |  | VRE |  |
|  | NL,<br>n/N with<br>data (%) | DK,<br>n/N with<br>data (%) | NL,<br>n/N with<br>data (%) | DK,<br>n/N with<br>data (%) | NL,<br>n/N with<br>data (%) | DK,<br>n/N with<br>data (%) | NL,<br>n/N with<br>data (%) | DK,<br>n/N with<br>data (%) |
| **Deceased** | **1/130 (1)** | **2/104 (2)** | **1/36 (3)** | **1/27 (4)** | **12/130 (9)** | **13/104 (12)** | **4/36 (11)** | **4/27 (15)** |
| **Censored<sup>b</sup>** |  |  |  |  | **1/130 (1)** |  |  |  |
| **Discharged** | **1/130 (1)** |  |  | **1/27 (4)** | **5/130 (4)** | **1/104 (1)** | **1/36 (3)** | **2/27 (7)** |
| **E. faecium-covering therapy** | **10/129 (8)** | **5/102 (5)** | **13/35 (37)** | **7/26 (27)** | **76/117 (65)** | **82/91 (90)** | **20/32 (62)** | **22/23 (96)** |
| - vancomycin iv | 9/129 (7) | 5/102 (5) | 12/35 (34) | 5/26 (19) | 72/117 (62) | 79/91 (87) | 8/32 (25) | 2/23 (9) |
| - linezolid iv |  |  | 1/35 (3) | 1/26 (4) | 1/117 (1) | 1/91 (1) | 6/32 (19) | 7/23 (30) |
| - daptomycin iv + linezolid iv |  |  |  |  |  |  |  | 10/23 (43) |
| - teicoplanin iv | 1/129 (1) |  |  |  | 2/117 (2) |  | 4/32 (12) |  |
| - linezolid po |  |  |  | 1/26 (4) |  |  | 2/32 (6) | 2/23 (9) |
| - daptomycin iv |  |  |  |  |  | 2/91 (2) |  |  |
| - vancomycin intrathecally |  |  |  |  | 1/117 (1) |  |  |  |
| - linezolid iv + tigecyclin iv |  |  |  |  |  |  |  | 1/23 (4) |
| **Appropriate therapy** | **10/129 (8)** | **5/102 (5)** | **1/35 (3)** | **2/26 (8)** | **76/117 (65)** | **82/91 (90)** | **12/32 (38)** | **20/23 (87)** |
| **Appropriate therapy or**<br>**central line removed** | **12/129 (9)** | **5/102 (5)** | **2/35 (6)** | **2/26 (8)** | **76/117 (65)** | **82/91 (90)** | **12/32 (38)** | **20/23 (87)** |

Abbreviations: DK, Denmark; iv, intravenously; NL, Netherlands; po, orally.
<sup>a</sup> Day 0 is the day of the index blood culture of the ARE/VRE bacteremia episode.
<sup>b</sup> One Dutch patient with ARE bacteremia was censored for the assessment of antibiotic therapy (not for mortality) due to transfer to another hospital.

Inclusion of inappropriate antibiotic therapy on the day of the index blood culture (day 0), in itself associated with mortality (RR 2.79 (95% CI 0.99-7.86)), did not alter the effect of VRE on 30 day mortality (Table 2). In models with an interaction between vancomycin resistance and appropriateness of therapy, VRE patients on inappropriate therapy increasingly fared worse over time compared to ARE patients on inappropriate therapy (Table 5). ARE patients on appropriate therapy had better survival than those on inappropriate therapy, but this protective effect seemed to diminish over time. The effect estimates for VRE patients on appropriate therapy were uncertain.

**Table 5.** Regression models for 30-day mortality evaluating appropriateness of therapy

|  | **Baseline after day ...** |  |  |
| --- | --- | --- | --- |
|  | **0** | **+1** | **+2** |
|  | **Unadjusted models** |  |  |
| *No of observations* | 289 | 283 | 274 |
| *No of events* | 95 | 88 | 80 |
| ARE – on inappropriate therapy | Reference | Reference | Reference |
| ARE – on appropriate therapy | 0.41 (0.10-1.74) | 0.82 (0.45-1.51) | 1.01 (0.57-1.79) |
| VRE – on inappropriate therapy | 1.24 (0.82-1.85) | 1.15 (0.71-1.88) | 1.40 (0.73-2.70) |
| VRE – on appropriate therapy | NA | 1.53 (0.68-3.46) | 1.32 (0.64-2.71) |
|  | **Adjusted models – main analysis** |  |  |
| *No of observations* | 289 | 276 | 268 |
| *No of events* | 95 | 87 | 80 |
| *No of variables* | 9 | 14 | 14 |
| *Events to variables* | 10.6 | 6.2 | 5.7 |
| ARE – on inappropriate therapy | Reference | Reference | Reference |
| ARE – on appropriate therapy | 0.33 (0.08-1.40) | 0.79 (0.43-1.45) | 0.82 (0.47-1.43) |
| VRE – on inappropriate therapy | 1.69 (1.09-2.61) | 2.01 (1.13-3.57) | 2.43 (0.94-6.33) |
| VRE – on appropriate therapy | NA | 5.79 (1.43-23.40) | 1.73 (0.83-3.61) |
|  | **Adjusted models – sensitivity analysis** |  |  |
| *No of observations* | 285 | 278 | 270 |
| *No of events* | 94 | 87 | 80 |
| *No of variables* | 16 | 17 | 15 |
| *Events to variables* | 5.9 | 5.1 | 5.3 |
| ARE – on inappropriate therapy | Reference | Reference | Reference |
| ARE – on appropriate therapy | 0.31 (0.08-1.13) | 0.69 (0.38-1.28) | 0.88 (0.42-1.84) |
| VRE – on inappropriate therapy | 1.42 (0.79-2.55) | 1.81 (0.99-3.31) | 2.38 (0.99-5.74) |
| VRE – on appropriate therapy | NA | 2.60 (0.55-12.32) | 2.12 (0.88-5.09) |

This table presents risk ratios (RR, with 95% CI) for 30-day mortality for the interaction between *vancomycin resistance* and *appropriateness of therapy*. The baseline for these models is positioned on three different moments, namely the end of the day of the index blood culture
(day 0), the end of the day after (day +1), and the end of the day thereafter (day +2). This implies that separately for each baseline, patients having died (or censored, as antibiotic therapy could not be fully assessed) before this moment were removed from the dataset.
*Appropriateness of therapy* refers to the antibiotic therapy provided at baseline. RRs are presented with and without adjustment for confounding variables. In the Supplementary Material, the adjustment procedure is described. All models include data from both the Netherlands and Denmark.

## Discussion

This study reveals that, after matching and further analytic control for confounders, VRE bacteremia was, compared to ARE bacteremia, associated with 54% higher risk for mortality after 30 days (RR 1.54, 95% CI 1.06-2.25). Yet, this increased risk of death must be explained by other factors than a delay in appropriate antibiotic therapy.

Our estimate for the effect of VRE on mortality is similar to the reported pooled OR of 2.52 (95% CI 1.9-3.4) in a meta-analysis from 2005 [12], which translates to a RR of 1.70 in case of 32% death rate in the non-exposed group [24], This seems remarkably identical, as the studies included in that meta-analysis had been performed before the availability of effective antibiotics for VRE, such as linezolid and daptomycin. Yet, a small study comparing older and newer antibiotics in 113 VRE bacteremias concluded that newer antibiotics had not brought discernable benefits to patient outcome [25], A later meta-analysis on the effect of VRE on mortality in the era of effective antibiotic therapy could only present an unadjusted estimate, and hence cannot be compared to our study [13]. Recent Australian and German studies all found point estimates reflecting increased mortality in case of vancomycin resistance, with confidence intervals indicating non-significance [26–28], By analyzing the *E. faecium* bacteremia subset and not including intermediate variables in the adjusted model, the OR of 1.283 (95% CI 0.801-2.057) from Kramer *et al*. most closely reflects our estimation procedure [27],

There are three causal pathways along which vancomycin resistance could lead to increased mortality: (i) increased virulence of VRE compared to ARE, (ii) less effective antibiotics for VRE than for ARE, and (iii) a delay in initiation of appropriate antibiotic therapy for VRE bacteremia. We cannot fully exclude a systematic difference in pathogenicity between ARE and VRE, as for example Bender *et al*. have shown that acquisition of *vanB* by *E. faecium* is accompanied by the transfer of larger genetic fragments [29], However, most studies conclude that both phenotypes belong to the same, well-characterized, core genomic lineage of *E. faecium* [4,7], Furthermore, to the best of our knowledge, there is no evidence that the appropriate antibiotic options for *E. faecium*, most prominently vancomycin, linezolid or daptomycin, have different efficacy for susceptible strains. In this study, vancomycin was mostly used for ARE, and linezolid and daptomycin for VRE.

The observed increased mortality in case of VRE bacteremia, therefore, could be expected to result from a delay in appropriate therapy, which has been implicated previously in worse outcomes in case of enterococcal bacteremia [30], However, our models that include appropriateness of therapy do not offer support for this hypothesis. VRE patients on inappropriate therapy continuously fare worse than ARE patients on inappropriate therapy. Therapy over time may be reflective of the evolving disease severity of the patient, and collider bias may be induced by conditioning on appropriateness of therapy [31]. This means that effect estimates of appropriateness of therapy after baseline may not reflect true causal associations. However, seeing that this trend is discernable from the day of the index blood culture onwards increases our confidence that the difference in duration until appropriate therapy is unable to explain the increased mortality in case of VRE bacteremia. A final indication for this stems from the comparison between countries in this study. Overall mortality in Denmark for both ARE and VRE bacteremia is higher than in Netherlands, although appropriate therapy for both types of bacteremia is initiated considerably faster in Denmark than in the Netherlands.

As these biologically plausible mediators cannot explain increased mortality due to vancomycin resistance, the possibility remains that these observed effects are due to unmeasured confounding. This possibility is supported by two additional observations. First, a measure for clinical disease severity immediately before bacteremia onset was not available for the Danish patients. For the Dutch patients, we could calculate the Acute Physiology score, and when included in our country-specific analyses, it substantially reduced the effect estimate for mortality (see Supplementary Material). Second, the association between vancomycin resistance and mortality persisted over the course of a full year. Infections may have long-term sequelae [32], but it seems unlikely that sustained mortality differences will emerge that can be causally related to vancomycin resistance. A Dutch population-based study reported that the incidence of recurrent bacteremia, an example of a long-term consequence, only marginally differed between ARE and VRE bacteremia [33], An alternative explanation is that underlying prognostic factors at the time of onset of enterococcal bacteremia were dissimilar.

Several limitations of this study should be discussed. First, results of this study may not apply to *E. faecium* bacteremia in general, as a non-random subset of ARE bacteremias was included. The matched design does not allow for direct comparisons of raw proportions other than for VRE vs. ARE. Second, some loss in precision may be expected in stratified analyses, as not all matched sets can be used for parameter estimation. Third, measurements of comorbidities and treatment restrictions differed between both countries, whereas these differences were not included in models. Fourth, duration until initiation of appropriate therapy could not be reliably measured in hours, and was reflected instead by calendar days.

Finally, some studies suggest that the incidence of infections with VRE occur on top of the existing incidence of infections caused by vancomycin-susceptible enterococci [34,35], In that case, a comparison between VRE bacteremia and an uninfected control group would be more appropriate, as described by Chiang *et al*. [36], Yet, it is important to note that these incidence rates may have been confounded by the fact that *E faecalis* was not separated from *E. faecium*, and that results from molecular epidemiological studies provided strong evidence that ARE and VRE occupy the same niche within the bacterial hospital ecology [4],

In conclusion, VRE bacteremia was, when compared to ARE bacteremia, associated with higher mortality. This could not be explained by delays in initiation of appropriate antibiotic therapy, although the relevant models are possibly underpowered and should be interpreted with caution. Because of the large heterogeneity among infected patients and the multiple determinants that mediate the outcome for patients developing *E. faecium* bacteremia, unmeasured confounding is a likely explanation. In that case, replacement of ARE infections by VRE infections would not lead to higher 30-day mortality. The alternative explanation is that VRE is more virulent than ARE. Given the resemblance of the core genomes of ARE and VRE, the genetic basis for hypervirulence would then be most likely encoded in the accessory genome, the mobilome. In that case, emergence of VRE could not only replace ARE infections but also increase the total burden of infection. Further studies are warranted to explore this possibility.

## Data Availability

Research data are not shared.

## Acknowledgment

We would like to thank all participating hospitals for making data collection possible. We specifically thank Marja Smid for her assistance with the data collection in the Netherlands, and Kim Gradel and Steen Rasmussen for extraction of comorbidity data in Denmark. We also thank Rob Willems for critically reviewing an earlier version of this manuscript.

*The Dutch VRE Bacteremia Investigators study group consists of*: Heidi S.M. Ammerlaan (Catharina Ziekenhuis, Eindhoven), Akke K. van der Bij (National Institute for Public Health and the Environment (RIVM), Bilthoven; present affiliation: Diakonessenhuis, Utrecht), Sybrandus N. Blank (Maxima Medisch Centrum, Eindhoven/Veldhoven), Marc J.M. Bonten (Universitair Medisch Centrum, Utrecht), Els I.G.B. de Brauwer (Zuyderland, Heerlen/Sittard-Geleen), Mirjam J.D. Dautzenberg (Universitair Medisch Centrum, Utrecht; present affiliation: Radboudumc, Nijmegen), Laura van Dommelen (Stichting PAMM, Veldhoven), Paul Gruteke (Onze Lieve Vrouwe Gasthuis, Amsterdam; Flevoziekenhuis, Almere), Rogier Jansen (Onze Lieve Vrouwe Gasthuis, Amsterdam), Saskia Kuipers (Radboudumc, Nijmegen), Marrigje H. Nabuurs-Franssen (Canisius-Wilhelmina Ziekenhuis, Nijmegen), Lieke Möller (Certe, Groningen), Jacobus M. Ossewaarde (Maasstad Ziekenhuis, Rotterdam), Wouter C. Rottier (Universitair Medisch Centrum, Utrecht), Wouter Rozemeijer (Noordwest Ziekenhuisgroep, Alkmaar), Gijs J.H.M. Ruijs (Isala, Zwolle), Annelot F. Schoffelen (National Institute for Public Health and the Environment (RIVM), Bilthoven), Fre W. Sebens (Deventer Ziekenhuis, Deventer; present affiliation: LabMicTA, Hengelo), Peter M. Schneeberger (Jeroen Bosch Ziekenhuis,’s-Hertogenbosch), Matthijs Tersmette (St. Antonius Ziekenhuis, Utrecht/Nieuwegein), Saara J. Vainio (VU medisch centrum, Amsterdam; present affiliation: St. Antonius Ziekenhuis, Utrecht/Nieuwegein), G. Paul Voorn (St. Antonius Ziekenhuis, Utrecht/Nieuwegein)

*The Danish Collaborative Bacteraemia Network (DACOBAN) study group consists of*: Magnus Arpi, Kim O. Gradel, Ulrich S. Jensen, Jenny D. Knudsen, Kristoffer Koch, Henrik C. Schønheyder, Mette Søgaard, Sara Thønnings, Jesper Smit, Mette Pinholt, Christian Ø. Andersen

Results from this study were presented at the 25th European Congress of Clinical Microbiology and Infectious Diseases, Copenhagen, Denmark (25-28 April 2015, oral presentation O090).

*Potential conflicts of interest*. All authors: No conflict.

## References

1. de Greeff SC, Mouton JW. NethMap 2017: consumption of antimicrobial agents and antimicrobial resistance among medically important bacteria in the Netherlands in 2016. Bergen, the Netherlands: Stichting Werkgroep Antibioticabeleid & Rijksinstituut voor Volksgezondheid en Milieu, 2017.

2. Borck Høg B, Korsgaard H, Sönksen UW. DANMAP 2016: use of antimicrobial agents and occurrence of antimicrobial resistance in bacteria from food animals, food and humans in Denmark. Lyngby & Copenhagen, Denmark: Statens Serum Institut, DTU Veterinærinstituttet & DTU Fødevareinstituttet, 2017.

3. Top J, Willems RJL, Bonten MJM. Emergence of CC17 Enterococcus faecium: from commensal to hospital-adapted pathogen. FEMS Immunol Med Microbiol 2008; 52: 297–308.

4. Guzman Prieto AM, van Schaik W, Rogers MRC, et al. Global emergence and dissemination of enterococci as nosocomial pathogens: Attack of the clones? Front Microbiol 2016; 7: 788.

5. Howden BP, Holt KE, Lam MMC, et al. Genomic insights to control the emergence of vancomycin-resistant enterococci. MBio 2013; 4:e00412–13.

6. Pinholt M, Gumpert H, Bayliss S, et al. Genomic analysis of 495 vancomycin-resistant Enterococcus faecium reveals broad dissemination of a vanA plasmid in more than 19 clones from Copenhagen, Denmark. J Antimicrob Chemother 2017; 72: 40–47.

7. Raven KE, Reuter S, Reynolds R, et al. A decade of genomic history for healthcare-associated Enterococcus faecium in the United Kingdom and Ireland. Genome Res 2016; 26: 1388–1396.

8. Bijzonder resistente micro-organismen (BRMO). Leiden, the Netherlands: Werkgroep Infectiepreventie, 2013.

9. Nationale infektionshygiejniske retningslinjer om supplerende forholdsregler ved infektioner og baerertilstand i sundhedssektoren (5. udgave). Copenhagen, Denmark: Central Enhed for Infektionhygiejne (Statens Serum Institut), 2016.

10. Johnstone J, Policarpio ME, Lam F, et al. Rates of blood cultures positive for vancomycin-resistant Enterococcus in Ontario: a quasi-experimental study. CMAJ Open 2017; 5:E273–E280.

11. Dik JWH, Dinkelacker AG, Vemer P, et al. Cost-analysis of seven nosocomial outbreaks in an academic hospital. PLoS One 2016; 11:e0149226.

12. DiazGranados CA, Zimmer SM, Klein M, Jernigan JA. Comparison of mortality associated with vancomycin-resistant and vancomycin-susceptible enterococcal bloodstream infections: a meta-analysis. Clin Infect Dis 2005; 41: 327–33.

13. Prematunge C, MacDougall C, Johnstone J, et al. VRE and VSE bacteremia outcomes in the era of effective VRE therapy: a systematic review and meta-analysis. Infect Control Hosp Epidemiol 2016; 37: 26–35.

14. Altorf-van der Kuil W, Schoffelen AF, de Greeff SC, et al. National laboratory-based surveillance system for antimicrobial resistance: a successful tool to support the control of antimicrobial resistance in the Netherlands. Euro Surveill 2017; 22:pii = 17–00062.

15. Gradel KO, Schønheyder HC, Arpi M, Knudsen JD, Østergaard C, Søgaard M. The Danish Collaborative Bacteraemia Network (DACOBAN) database. Clin Epidemiol 2014; 6: 301–308.

16. Cummings P. Re: ‘Estimating the relative risk in cohort studies and clinical trials of common outcomes’. Am J Epidemiol 2004; 159: 213.

17. Greenland S, Daniel R, Pearce N. Outcome modelling strategies in epidemiology: traditional methods and basic alternatives. Int J Epidemiol 2016; 45: 565–575.

18. R Core Team. R: A language and environment for statistical computing. Vienna, Austria: 2020.

19. Therneau T. A package for survival analysis in R (R package version 3.1–11). 2020;

20. Gray B. cmprsk: subdistribution analysis of competing risks (R package version 2.2–9). 2019;

21. Harrell Jr FE. rms: Regression Modeling Strategies (R package version 5.1–4). 2019;

22. Buuren S van, Groothuis-Oudshoorn K. mice: multivariate imputation by chained equations in R. J Stat Softw 2011; 45.

23. Dahl DB, Scott D, Roosen C, Magnusson A, Swinton J. xtable: export tables to LaTeX or HTML (R package version 1.8–4). 2019;

24. Zhang J, Yu KF. What’s the relative risk? A method of correcting the odds ratio in cohort studies of common outcomes. JAMA 1998; 280: 1690–1.

25. Erlandson KM, Sun J, Iwen PC, Rupp ME. Impact of the more-potent antibiotics quinupristin-dalfopristin and linezolid on outcome measure of patients with vancomycin-resistant Enterococcus bacteremia. Clin Infect Dis 2008; 46: 30–6.

26. Cheah ALY, Spelman T, Liew D, et al. Enterococcal bacteraemia: factors influencing mortality, length of stay and costs of hospitalization. Clin Microbiol Infect 2013; 19:E181–E189.

27. Kramer TS, Remschmidt C, Werner S, et al. The importance of adjusting for enterococcus species when assessing the burden of vancomycin resistance: a cohort study including over 1000 cases of enterococcal bloodstream infections. Antimicrob Resist Infect Control 2018; 7: 133.

28. Dubler S, Lenz M, Zimmermann S, et al. Does vancomycin resistance increase mortality in Enterococcus faecium bacteraemia after orthotopic liver transplantation? A retrospective study. Antimicrob Resist Infect Control 2020; 9: 22.

29. Bender JK, Kalmbach A, Fleige C, Klare I, Fuchs S, Werner G. Population structure and acquisition of the vanB resistance determinant in German clinical isolates of Enterococcus faecium ST192. Sci Rep 2016; 6: 21847.

30. Zasowski EJ, Claeys KC, Lagnf AM, Davis SL, Rybak MJ. Time is of the essence: the impact of delayed antibiotic therapy on patient outcomes in hospital-onset enterococcal bloodstream infections. Clin Infect Dis 2016; 62: 1242–1250.

31. Hernán MA, Hernández-Díaz S, Robins JM. A structural approach to selection bias. Epidemiology 2004; 15: 615–25.

32. Leibovici L. Long-term consequences of severe infections. Clin Microbiol Infect 2013; 19: 510–512.

33. Woudt SHS, de Greeff SC, Schoffelen AF, Vlek ALM, Bonten MJM, Infectious Diseases Surveillance Information System-Antimicrobial Resistance (ISIS-AR) Study Group. Antibiotic resistance and the risk of recurrent bacteremia. Clin Infect Dis 2018; 66: 1651–1657.

34. Ammerlaan HSM, Harbarth S, Buiting AGM, et al. Secular trends in nosocomial bloodstream infections: antibiotic-resistant bacteria increase the total burden of infection. Clin Infect Dis 2013; 56: 798–805.

35. Popiel KY, Miller MA. Evaluation of vancomycin-resistant enterococci (VRE)-associated morbidity following relaxation of VRE screening and isolation precautions in a tertiary care hospital. Infect Control Hosp Epidemiol 2014; 35: 818–25.

36. Chiang H-Y, Perencevich EN, Nair R, et al. Incidence and outcomes associated with infections caused by vancomycin-resistant enterococci in the United States: systematic literature review and meta-analysis. Infect Control Hosp Epidemiol 2017; 38: 203–215.

